# Longitudinal glioma monitoring via cerebrospinal fluid cell-free DNA: one patient at a time

**DOI:** 10.1101/2024.02.21.24303164

**Authors:** Cecile Riviere-Cazaux, Xiaoxi Dong, Wei Mo, Chao Dai, Lucas P. Carlstrom, Amanda Munoz-Casabella, Rahul Kumar, Keyvan Ghadimi, Cody L. Nesvick, Katherine M. Andersen, Matthew D. Hoplin, Nicholas Canaday, Ignacio Jusue-Torres, Noor Malik, Jian L. Campian, Michael W. Ruff, Joon H. Uhm, Jeanette E. Eckel Passow, Timothy J. Kaufmann, David M. Routman, Sani H. Kizilbash, Arthur E. Warrington, Robert B. Jenkins, Pan Du, Shidong Jia, Terry C. Burns

**Affiliations:** Departments of Neurological Surgery, Mayo Clinic, Rochester, MN, USA; Predicine, Inc., Hayward, CA, USA; Department of Neurological Surgery, The Ohio State University, Columbus, OH, USA; Leo M. Davidoff Department of Neurological Surgery, Albert Einstein College of Medicine, Bronx, NY, USA; Departments of Medical Oncology, Mayo Clinic, Rochester, MN, USA; Departments of Neurology, Mayo Clinic, Rochester, MN, USA; Departments of Quantitative Health Sciences, Mayo Clinic, Rochester, MN, USA; Departments of Radiology, Mayo Clinic, Rochester, MN, USA; Departments of Radiation Oncology, Mayo Clinic, Rochester, MN, USA; Departments of Laboratory Medicine and Pathology, Mayo Clinic, Rochester, MN, USA

**Keywords:** glioma, biomarker, cerebrospinal fluid, monitoring, cell-free DNA

## Abstract

**IMPORTANCE:** Current methods for glioma treatment response assessment are limited. Intracranial cerebrospinal fluid (CSF) may provide a previously untapped source of longitudinal biomarkers, such as cell-free DNA (cfDNA), for disease monitoring.

**OBJECTIVE:** To assess the feasibility of obtaining longitudinal intracranial CSF cfDNA from patients with gliomas during their disease course.

**DESIGN:** This case series was initiated in 2021, with patients followed until last clinical follow-up (death or present time).

**SETTING:** This single-center study was conducted at a large academic medical center.

**PARTICIPANTS:** Adults with gliomas were recruited for longitudinal intracranial CSF collection using 1) Ommaya reservoirs, from which CSF would be sampled on at least two separate occasions, or 2) CSF collection from other clinically indicated CSF access devices, such as ventriculoperitoneal (VP) shunts.

**INTERVENTIONS:** CSF was collected from Ommaya reservoirs in four patients and from an existing VP shunt in one patient.

**MAIN OUTCOMES AND MEASURES:** The study aimed to collect CSF for biobanking and biomarker discovery, with the hypothesis that CSF could serve as a source of longitudinally acquirable biomarkers.

**RESULTS:** Five patients (2 females, 3 males; median: 40 years, range 32-64 years) underwent longitudinal intracranial CSF collection via Ommaya reservoirs (n=4/5 patients) or VP shunt (n=1/5). Three patients had glioblastoma and two had astrocytoma, IDH-mutant, grade 4. In total, thirty-five CSF samples were obtained (median: 3.80 mL; 0.5-5 mL), with 30 (85.7%) yielding sufficient cfDNA for Next-Generation Sequencing (n=28) or Low-Pass Whole Genome sequencing (all samples). Tumor fraction was found to increase with radiographic progression. Changes in variant allelic frequencies (VAFs) may be seen within individual patients after resection and chemoradiation. In two patients, changes in tumor-specific IDH1 VAF correlated with CSF D-2-hydroxyglutarate levels, the oncometabolite of IDH mutant tumors. Copy number burden (CNB) decreased below the limit of quantification during treatment.

**CONCLUSIONS AND RELEVANCE:** Longitudinal CSF cfDNA can feasibly be obtained via CSF access devices in patients with gliomas during their disease course. Ongoing studies will evaluate hypotheses generated in this case series regarding how longitudinal CSF cfDNA could be utilized to sensitively detect changes in disease burden.

**Trial Registration:** NCT04692324 https://clinicaltrials.gov/study/NCT04692324; NCT04692337 https://clinicaltrials.gov/study/NCT04692337

**QUESTION:** What is the feasibility of obtaining longitudinal intracranial cerebrospinal fluid cell-free DNA from patients with high-grade gliomas to evaluate changes during treatment?

**FINDING:** In this case series, we find that CSF cfDNA can feasibly be obtained throughout treatment via CSF access devices. We find that changes in tumor fraction or tumor-associated variant allele frequencies (VAFs) may correlate with disease trajectory, with VAFs positively correlating to other tumor-associated candidate biomarkers.

**MEANING:** Longitudinal cerebrospinal cell-free DNA may inform the impact of treatment throughout a specific patient’s disease course, from the time of resection through radiographic progression.

## INTRODUCTION

Few tools exist to monitor glioma burden and biological response to standard-of-care chemoradiation or experimental therapies. The availability of serial biopsy samples has enabled rapid progress in monitoring for some cancers, including breast cancer, melanoma, and blood-based cancers^1–5^. However, the requirement for brain biopsy to obtain glioma tissue during treatment has hindered monitoring^6, 7^. MRI is standard-of-care for treatment response assessment, but its sensitivity and specificity remain a limitation^8, 9^. Based on well-established cancer genomic markers, cell-free DNA (cfDNA) in liquid biopsies has become of growing interest for cancer detection and monitoring, particularly in plasma^10–13^. Numerous trials are now leveraging cfDNA to guide therapeutic decision making^14–19^. However, the blood-brain barrier has hampered detection of tumor-associated cfDNA in plasma, also known as cell-tumor DNA (ctDNA), with studies reporting a low detection rate, often between 10-50%^20–22^.

Studies in other cancers, such as prostate cancer^23, 24^, demonstrate that proximal fluids improve detection of tumor-derived biomarkers^25^. Similarly, as the liquid source closest to the tumor, cfDNA detection rates in cerebrospinal fluid (CSF) from patients with brain tumors, including gliomas and brain metastases, increase to an average of approximately 85% in more recent studies^26–30^. CSF cfDNA studies in glioma have repeatedly demonstrated the superiority of CSF to plasma for cfDNA detection^27, 29^, as well as the fidelity of cfDNA for reporting mutations detected in tissue^31^. Instead of targeted sequencing for mutations, recent studies have increasingly utilized Next-Generation Sequencing (NGS) or low-pass whole genome sequencing (LP-WGS) to characterize the global genomic landscape of glioma CSF cfDNA for tumor diagnosis and identification of treatment-resistant mutations^32, 33^.

Longitudinal CSF cfDNA may be useful for disease monitoring. A previous study reported on changes in tumor-associated mean allele frequencies between two CSF timepoints^27^. However, to our knowledge, no study has obtained longitudinal intracranial CSF samples for cfDNA analysis during a patient’s disease course. We hypothesized that intracranial CSF could be an abundant source of longitudinally acquirable cfDNA. Herein, we report our initial experience utilizing CSF access devices to determine how CSF cfDNA changes during treatment in patients with glioblastoma (GBM) or astrocytoma, IDH mutant (CNS WHO grade 4). Moreover, we identify remaining questions to guide ongoing and future studies aiming to utilize CSF cfDNA for disease burden monitoring during standard-of-care or clinical trials for patients with gliomas.

## MATERIALS AND METHODS

### Patient cohort

Informed consent was provided by each patient for collection of CSF in the operating room or clinic under biobanking, CSF biomarkers (NCT04692324, to obtain CSF from clinically indicated CSF devices), or Ommaya reservoir (NCT04692337, for research) protocols, each of which were approved via the Mayo Clinic Institutional Review Board. For all studies, inclusion criteria included adult patients with known or suspected brain neoplasms. For NCT04692337, inclusion criteria also included planned resection of the neoplasm and willingness to have the Ommaya sampled on 2 future occasions. Exclusion criteria for NCT04692324 included a clinical contraindication to the intended route of CSF access, and for NCT04692337, any prior wound infection or contraindication to the Ommaya.

### CSF collection

Patients had CSF access devices for longitudinal CSF access (4 research Ommaya reservoirs, 1 clinical ventriculoperitoneal shunt). The ventricular catheters were placed into the resection cavity, which contacted the ventricular system in each patient. CSF was sampled intra-operatively from the surgical field. In follow-up cases, 20 mL of CSF was slowly withdrawn from the Ommaya reservoir or shunt. CSF was maintained on ice or at 4°C (typically <1 hour), then centrifuged at 400G for 10 minutes at 4°C. De-identified aliquots were stored at -80°C in cryovials. Samples were annotated to include the known pathology, date of collection, and ongoing treatment, as well as technical variables including sample color, collection, and processing times.

### Cell-free DNA extraction and sequencing

Thirty-five CSF samples were analyzed across the 5 patients (median number of samples per patient: 7 (range: 3-12); median volume per sample: 3.80 mL (range: 0.5-5 mL)), of which 30 (85.7%) yielded sufficient cfDNA for testing. Patient samples were analyzed using two comprehensive NGS platforms, PredicineCARE and PredicineSCORE (Predicine, Inc.), to generate genomic aberration profiles and derive genome-wide copy number burden (CNB) score. Briefly, cell-free DNA (cfDNA) were subjected to library construction. PredicineCARE targeted hybridization enrichment assay was used to generate the landscape of genomic alterations including single-nucleotide variants (SNV), insertions and deletions (indels), CNVs, and gene fusions^34–37^. PredicineSCORE low pass whole genome sequencing assay was used to generate copy number profiles and genome-wide copy number burden (CNB) score, which represents a comprehensive genome-wide measure of copy number status, including amplifications and deletions if any, across the entire chromosome arms. See Supplemental methods for further details.

## RESULTS

### Patient 24 – Recurrent glioblastoma

The first patient to enroll on our Ommaya reservoir trial (Patient 24 in our CSF biobank) was a female in her late 50s with suspected recurrence of her GBM after standard chemoradiation (**Fig. 1A**). At her first post-operative CSF sample on post-operative day (POD) 26 after re-resection, a new region of substantial periventricular progression was observed on MRI, prompting initiation of lomustine (CCNU). At her next sample on POD75, further progression was noted, prompting transition to bevacizumab. The final CSF sample was obtained on POD118, at which time MRI showed ongoing progression on bevacizumab. She transitioned to hospice and passed away on POD152.

**Figure 1.**
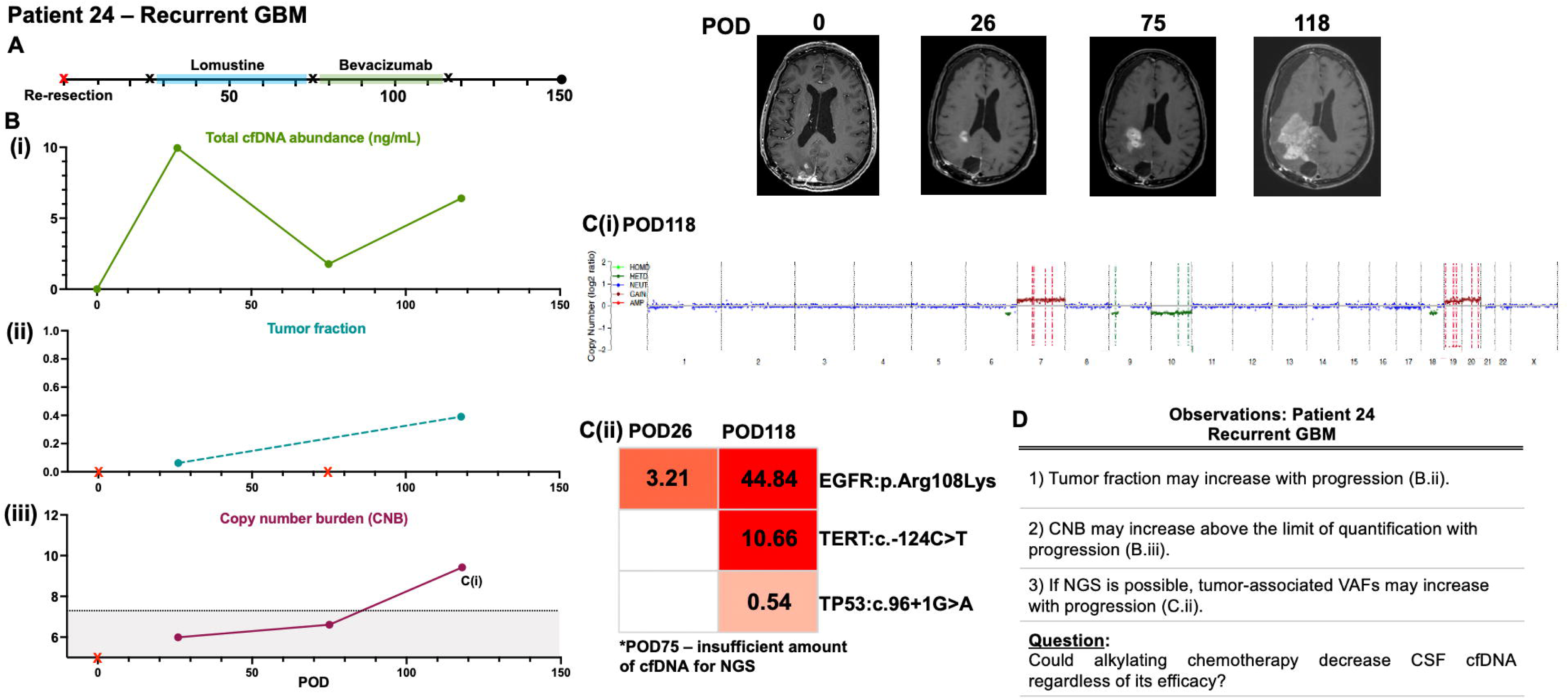
Longitudinal cerebrospinal fluid cell-free DNA from patient 24 (recurrent glioblastoma). **(A)** The timeline of the clinical course of patient 24 is depicted starting from post-operative (POD) 0, the day of resection for her recurrent GBM. Black X indicates when CSF samples were obtained and sequenced; red X indicates when CSF was sampled but had insufficient cfDNA for analysis. Correlative MRIs are shown for each timepoint. **(B) (i)** Total cell-free (cfDNA) abundance, **(ii)** tumor fraction, and **(iii)** copy number burden (CNB) were calculated from each CSF sample. CNB<7.5 (grey box) is below the limit of quantification. **(C)(i)** Copy number plots were generated for each CSF sample; POD118 is shown, and **(ii)** The variant allele frequencies (VAFs) were calculated for the two samples where sufficient cfDNA was obtained for next-generation sequencing (NGS), at POD26 and 118. **(D)** Observations and questions were raised by patient 24’s data. Red X = CSF obtained, insufficient cfDNA for testing.

Ventricular CSF was sent from the surgical field and was found to contain insufficient cfDNA (0.00 ng/mL) for baseline testing (**Fig. 1Bi)**. By contrast, the POD26 sample contained 9.95 ng/mL that decreased to 1.77 ng/mL after lomustine (POD75), despite radiographic disease progression after lomustine. This sample was insufficient for NGS, hampering evaluation of tumor fraction that otherwise increased 6.28x between POD26 and POD118 (**Fig. 1Bii)**. LP-WGS was also performed and revealed that copy number burden (CNB) increased above the limit of quantification by POD118 (**Fig. 1Biii**). Trisomy 7, loss of 9p (including CDKN2A/B), and gains on chromosomes 19 and 20 were detectable on the copy number plot by POD118 (**Fig. 1Ci)**.

Consistent with the patient’s known EGFR-amplified tumor, the variant allele frequency (VAF) for mutant EGFR increased from 3.21% to 44.84% between her pre-lomustine and post-bevacizumab samples (**Fig. 1cii).** The patient’s glioma harbored a TERT C228T mutation that was detectable at a VAF of 10.66% by the post-bevacizumab sample (POD118).

Initial observations generated by patient 24 (**Fig. 1D)** included that tumor fraction and VAFs may increase with progression (**Fig. 1Bii, 1Cii)**. CNB may also increase above the estimated limit of quantification (CNB>7.5) (**Fig. 1Biii).** However, despite radiographic progression at POD75, few data points were available due to limited cfDNA abundance after lomustine. This raised the question of whether alkylating therapy may decrease CSF cfDNA, even if ineffective. With these initial observations, we sought to evaluate patients during therapy from the time of original diagnosis.

### Patient 78 – Glioblastoma

Patient 78 (male in early 60s) underwent Ommaya placement during resection of a cystic lesion that proved to be GBM (**Fig. 2A**). The patient enrolled in a trial of pembrolizumab with standard chemoradiation. Imaging raised concern for progression around POD270, prompting initiation of regorafenib, a tyrosine kinase inhibitor. Further radiographic progression prompted transition to bevacizumab, with radiographic response. The patient transitioned to hospice and passed away on POD729.

**Figure 2.**
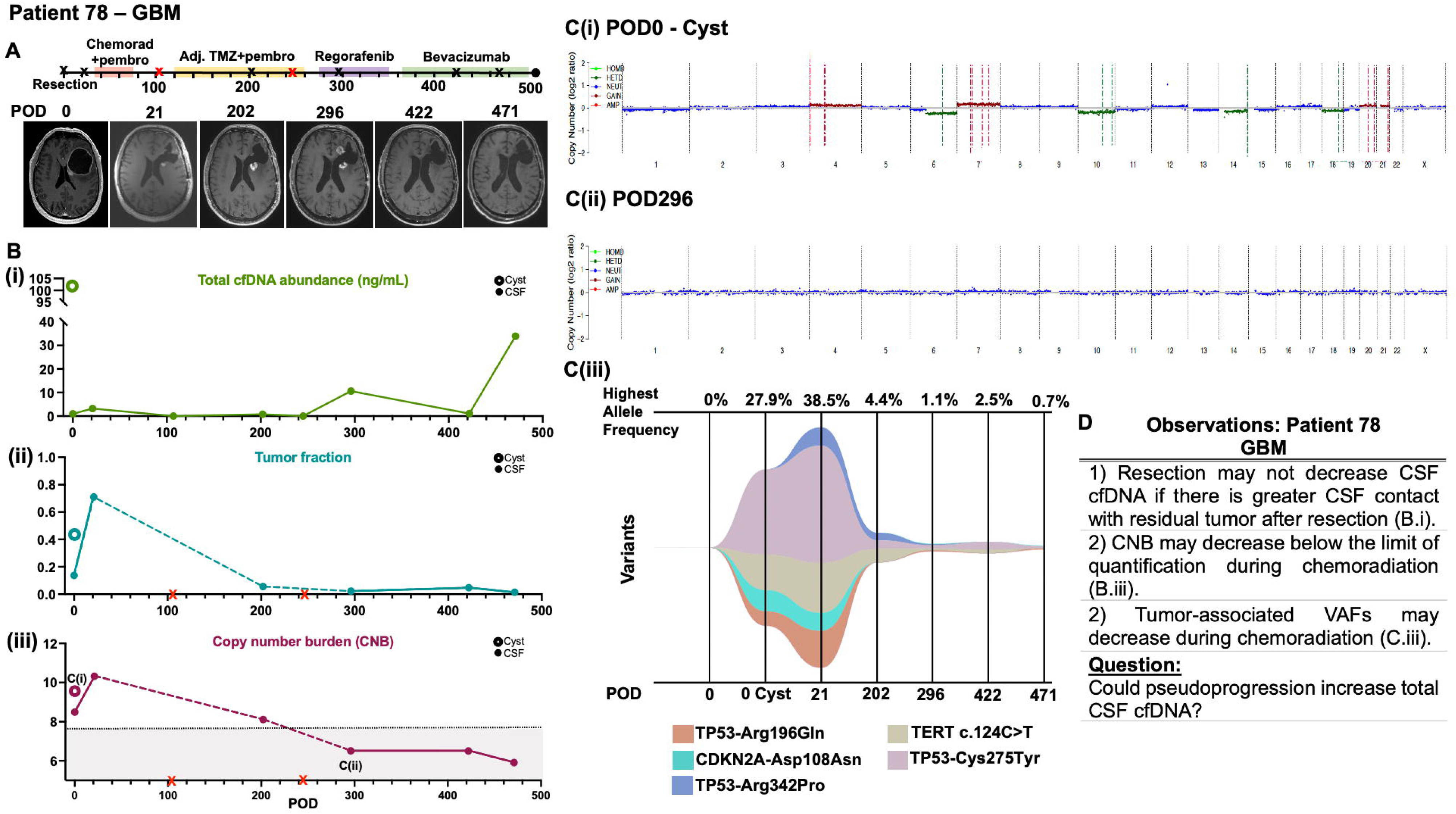
Longitudinal cerebrospinal fluid cell-free DNA from patient 78 (glioblastoma). **(A)** The timeline of the clinical course of patient 78 is depicted starting from post-operative (POD) 0, the day of resection for his primary GBM. On POD0, both CSF (solid circle) and cyst fluid (hollow circle) were analyzed. The patient underwent treatment with pembrolizumab in addition to standard chemoradiation. Black X indicates when CSF samples were obtained and sequenced; red X indicates when CSF was sampled but had insufficient cfDNA for analysis. Correlative MRIs are shown for each timepoint. **(B)(i)** Total cell-free DNA (cfDNA) abundance, **(ii)** tumor fraction, and **(iii)** copy number burden (CNB) were calculated from each CSF sample. **(C)(i-ii)** Copy number plots were generated for each sample; POD0 from the cyst fluid and POD202 are shown. **(iii)** Fishplot depicting the variant allele frequency of five detected mutations from POD0 to 471. **(D)** Observations and questions were raised by patient 78’s data. Red X = CSF obtained, insufficient cfDNA for testing.

CSF from a cortical sulcus and cyst fluid were both obtained during surgery, the latter of which yielded 104.2x more cfDNA (**Fig. 2Bi**). Total cfDNA abundance, tumor fraction, and CNB increased in post-resection CSF relative to the sulcal CSF sample (**Fig. 2Bi-iii**), likely reflecting the combined effect of recent tissue disruption and increased tumor contact with post-operatively sampled ventricular CSF. These measures then decreased with chemoradiation and pembrolizumab (POD202 versus 21; **Fig. 2Bi-iii**). No CSF cfDNA was extractable at POD107 immediately after chemoradiation. Despite suspected radiographic progression around POD270 which prompted initiation of regorafenib, no cfDNA could be extracted from CSF obtained at POD245. CSF obtained during regorafenib treatment demonstrated increased total cfDNA abundance, with CNB decreasing below the limit of quantification (POD 296; **Fig. 2Bi; ii**). cfDNA abundance declined with initiation of bevacizumab prior to increasing again by POD471, suggesting relative variability in cfDNA abundance despite minimal radiographic changes (**Fig 2Bii).** In contrast, CNB remained below the limit of quantification (**Fig. 2Biii**). +7, -10, +19 and +20 and other alterations were observed on the copy number plots on POD0 and could not be detected after chemoradiation and pembrolizumab (**Fig. 2Ci-ii**). TP53 and TERT VAFs initially increased after bevacizumab prior to decreasing by POD471 (**Fig. 2Ciii; Supplemental Fig. 1A)**. In sum, initial observations for patient 78 (**Fig. 2D)** included that resection may not decrease CSF cfDNA even by POD21 (**Fig. 2Bi).** CNB and tumor-associated VAFs may decrease during chemoradiation (**Fig. 2Biii; Ciii)**. Increased radiographic enhancement, but decreased CNB and VAFs, at POD296 prompted the question of whether pseudoprogression could increase total CSF cfDNA.

### Patient 79 – Glioblastoma

Patient 79 is a female in her early 30s who underwent resection for a GBM (**Fig. 3A)**. Given an expected subtotal resection and the highly vascular nature of her tumor, an EVD was also placed at the time of surgery in addition to her Ommaya reservoir. MSH2 and MSH6 staining revealed loss of expression, diagnostic of defective DNA mismatch repair. Next Generation Sequencing revealed over 200 mutations supportive of a hypermutant phenotype. Considering literature support for immunotherapy in hypermutation phenotypes^38^, she was treated with pembrolizumab, which began prior to chemoradiation. Radiographic progression near POD300 resulted in discontinuation of pembrolizumab and initiation of bevacizumab to which there was radiographic response.

**Figure 3.**
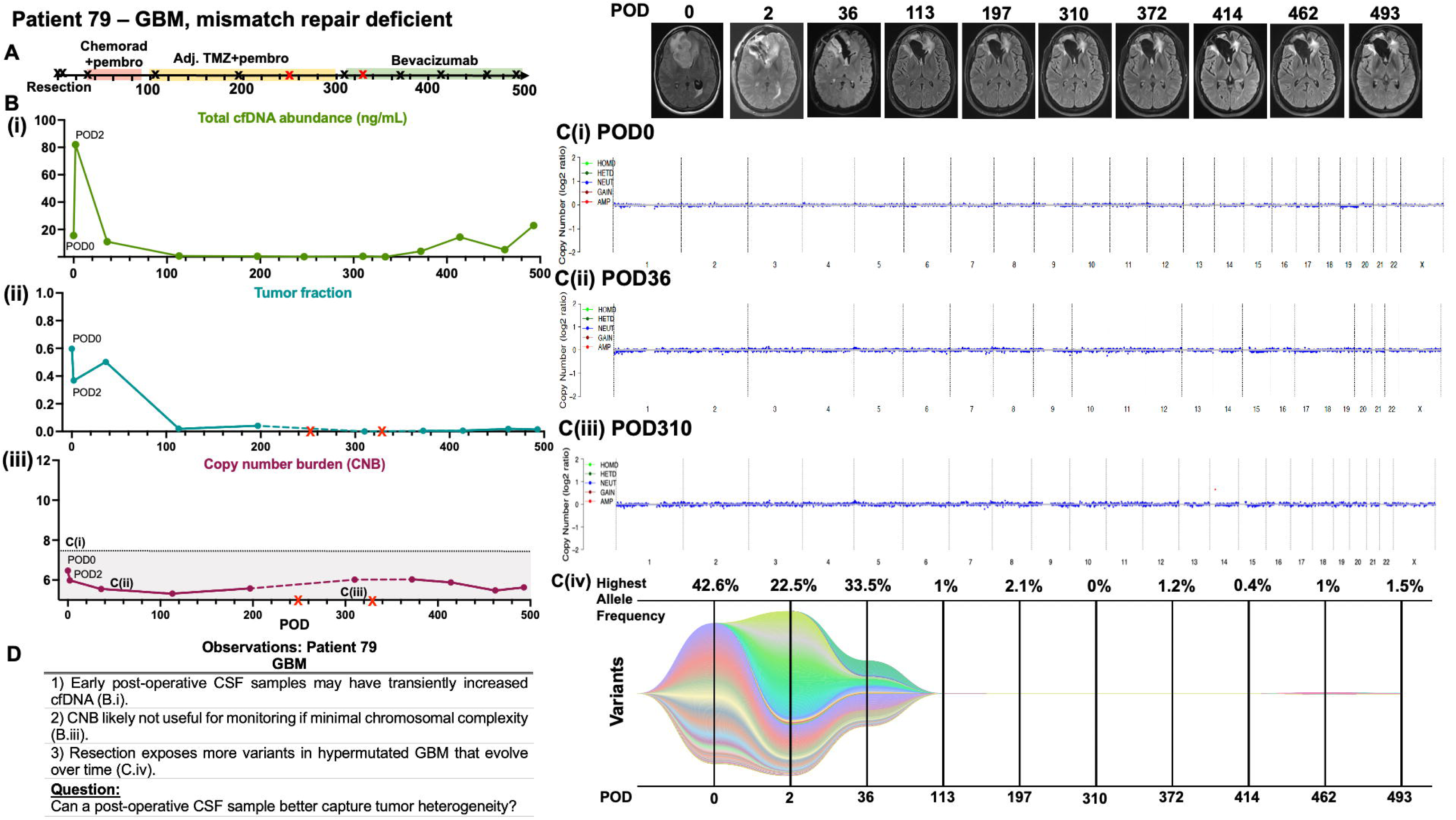
Longitudinal cerebrospinal fluid cell-free DNA from patient 79 (glioblastoma, mismatch repair deficient). **(A)** The timeline of the clinical course of patient 79 is depicted starting from post-operative (POD) 0, the day of resection for her glioblastoma that was mismatch repair deficient. She also underwent treatment with pembrolizumab in addition to standard chemoradiation. Black X indicates when CSF samples were obtained and sequenced; red X indicates when CSF was sampled but had insufficient cfDNA for analysis. Correlative MRIs are shown for each timepoint. Dashed lines = missing data between two points. **(B)(i)** Total cell-free DNA (cfDNA) abundance, **(ii)** tumor fraction, and **(iii)** copy number burden (CNB) were calculated from each CSF sample. **(C)(i-iii)** Copy number plots were generated for each CSF sample; POD0, 36, and 310 are shown, and **(iv**) fishplot depicting the variant allele frequency of numerous detected mutations from POD0 to 493. **(D)** Observations and questions were raised by patient 79’s data. Red X = CSF obtained, insufficient cfDNA for testing.

Intra-operatively obtained CSF yielded 15.60 ng/mL of cfDNA that transiently increased to 82.00 ng/mL when CSF was obtained from the EVD on POD2 (**Fig. 3Bi)**. In contrast, tumor fraction decreased from 0.597 to 0.368 (**Fig. 3Bii)**, potentially suggesting that much of the cfDNA on POD2 was from post-operative tissue disruption. Interestingly, despite the hypermutated phenotype, this tumor lacked evidence of chromosomal complexity, with a stably low CNB below the limit of quantification and no obvious chromosomal alterations on the whole genome plots (**Fig. 3Biii; 4Ci-iii**), unlike most GBMs that harbor +7/-10q alterations. In stark contrast, the longitudinal variant allele frequency plot demonstrated marked evolution of mutations over time (**Fig. 3Civ)**. POD2 revealed variants not seen on POD0, likely reflecting incomplete sampling of a heterogeneous lesion in the initial CSF. New variants seen on POD36 likely represent ongoing disease evolution prior to chemotherapy.

In summary (**Fig. 3D)**, samples from patient 79 suggested that early post-operative samples may have transiently increased cfDNA (**Fig. 3Bi**). Resection may expose more variants in a heterogeneous tumor, that may then evolve further prior to treatment (**Fig. 3Civ**). Given these findings, this patient’s data raised the question of whether a post-operative CSF sample may better sample or capture tumor heterogeneity (**Fig. 3D**).

### Patient 98 – Astrocytoma, IDH mutant (CNS WHO grade 4)

Patient 98 is a male in his early 40s who underwent resection of an astrocytoma, IDH mutant (CNS WHO grade 4) and Ommaya placement (**Fig. 4A)**. In addition to standard-of-care chemoradiation, he participated in an immunotherapy trial wherein he was randomized to either NT-I7 or placebo at an outside facility. No post-operative CSF samples were obtained until POD146 after chemoradiation when MRI raised concern for progression versus pseudoprogression.

**Figure 4.**
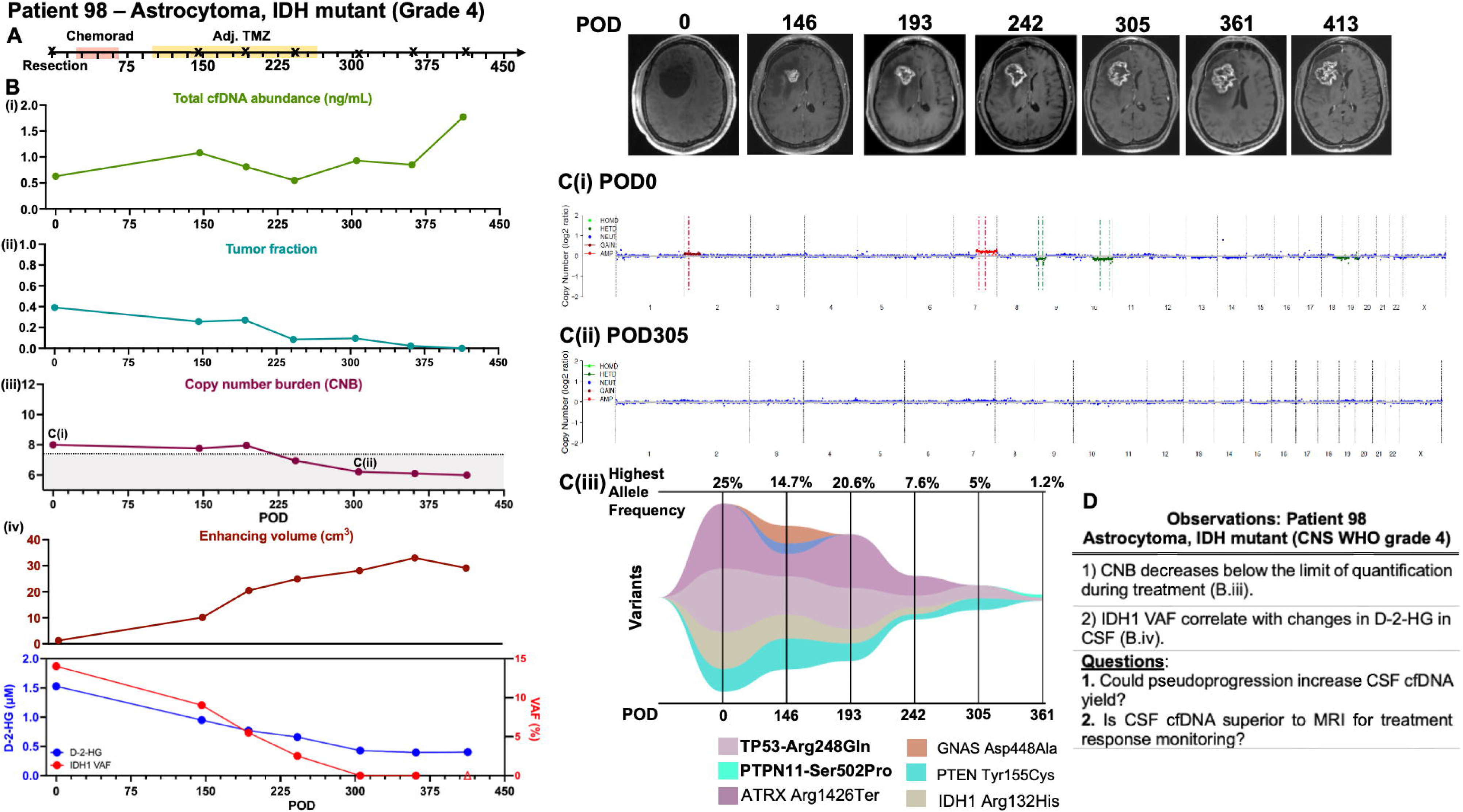
Longitudinal cerebrospinal fluid cell-free DNA from patient 98 (astrocytoma, IDH mutant, CNS WHO grade 4). **(A)** The timeline of the clinical course of patient 98 is depicted starting from post-operative (POD) 0, the day of resection for his astrocytoma, IDH mutant, CNS WHO grade 4. Black X indicates when CSF samples were obtained and sequenced; red X indicates when CSF was sampled but had insufficient cfDNA for analysis. **(B) (i)** Total cell-free DNA (cfDNA) abundance, **(ii)** tumor fraction, and **(iii)** copy number burden (CNB) were calculated from each CSF sample. Additionally, **(iv)** volumetrics was performed to calculate the enhancing tumor volume (cm^3^) from each timepoint. The variant allele frequency (VAF) for the IDH1 mutation and the levels of D-2-hydroxyglutarate (D-2-HG, in µM) were also quantified at each timepoint. Hollow triangle: no IDH1 VAF detected. **(C)(i-ii)** Copy number plots were generated for each CSF sample; POD0 and 305 are shown. **(iii)** Fishplot depicting the VAFs of six detected mutations from POD0 to 361. **(D)** Observations and questions were raised by patient 98’s data.

Despite variable changes in cfDNA abundance and increasing radiographic contrast enhancement, tumor fraction decreased and CNB fell below the limit of quantification during treatment and observation (**Fig. 4Bi-iii)**. Indeed, while contrast-enhancing volume increased, decreasing IDH1 VAF also correlated with decreasing CSF D-2-hydroxyglutarate (D-2-HG) levels, an oncometabolite of IDH mutant tumors (**Fig. 4Biv**)^39^. Gain of 2p, gain of 7q, loss of 9p (without CDKN2A/B homozygous deletion), and loss of 10q, all common alterations in astrocytoma, IDH-mutant, were detected in the POD0 sample, but these were no longer detectable by POD305 (**Fig. 4Ci-ii).** In addition to IDH1 (**Fig. 4Biv**), VAFs for most detected mutations decreased with treatment, with only one of the originally detected variants present by POD361 when the patient was on observation (**Fig. 4Ciii; Supplemental Fig. 1B**).

In summary (**Fig. 4D)**, observations from patient 98 suggested that CNB may decrease below the limit of quantification with treatment (**Fig. 4Biii)**, as seen with patient 78 (**Fig. 2Biii**). Moreover, IDH1 VAF may correlate with changes in CSF D-2-HG (**Fig 4Biv)**. As with patient 78, we again questioned whether pseudoprogression could increase CSF cfDNA yield. Agreement between tumor fraction, IDH1 VAF, 2-HG, and CNB despite increasing enhancing volume prompted the hypothesis that CSF cfDNA may be superior to MRI for treatment response monitoring.

### Patient 138 – Astrocytoma, IDH mutant (CNS WHO grade 4)

Patient 138 is a male in his early 30s who underwent pre-operative EVD placement due to clinical decline with ventriculomegaly. His EVD was maintained after tumor resection, which revealed an astrocytoma, IDH mutant (CNS WHO grade 4). Due to difficulty weaning the EVD, a ventriculoperitoneal shunt was placed (**Fig. 5A**). By POD214, radiographic disease progression was suspected, and the patient was initiated on lomustine.

**Figure 5.**
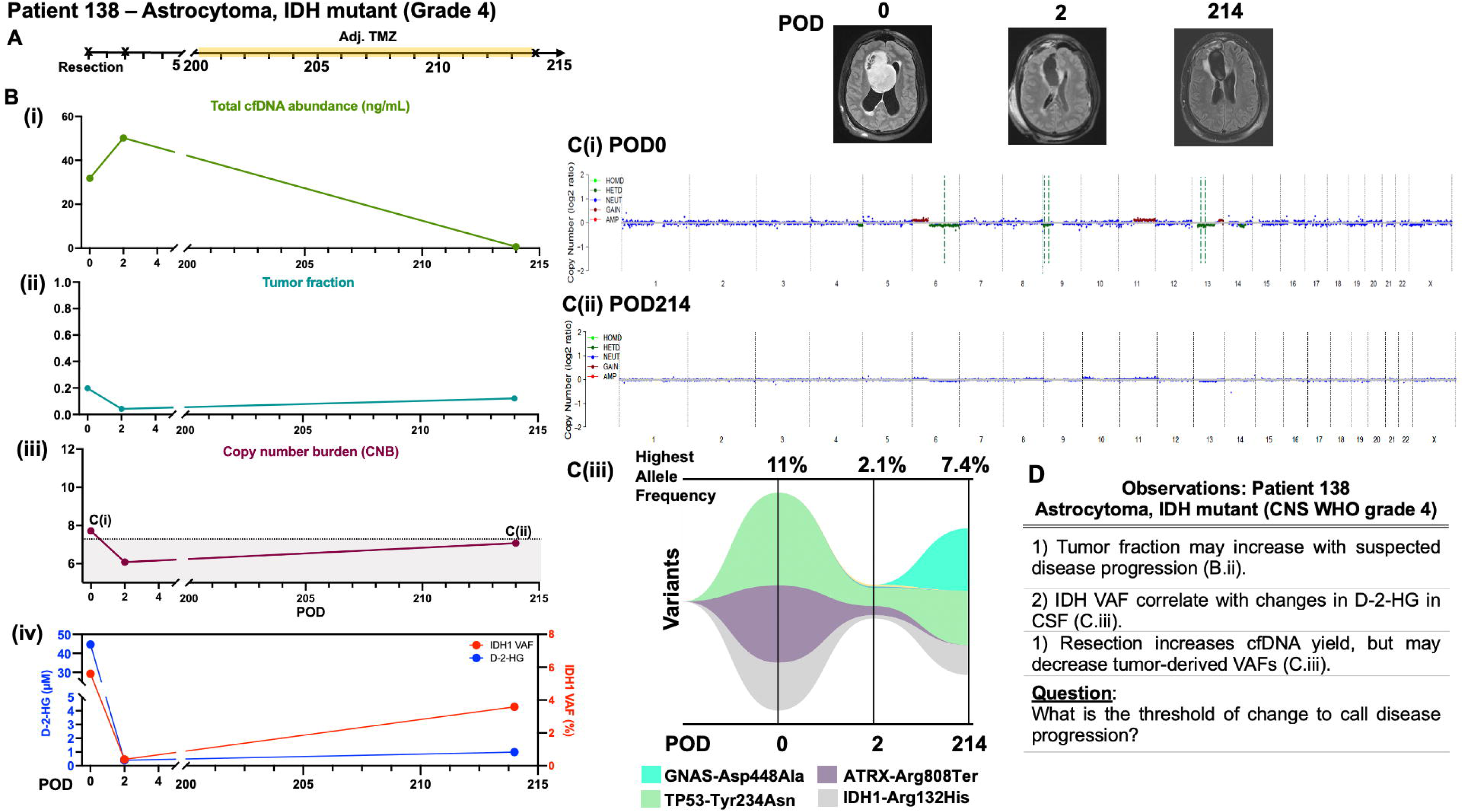
Longitudinal cerebrospinal fluid cell-free DNA from patient 138 (astrocytoma, IDH mutant, grade 4). **(A)** The timeline of the clinical course of patient 138 is depicted starting from post-operative (POD) 0, the day of resection for his astrocytoma, IDH mutant, grade 4. Black X indicates when CSF samples were obtained and sequenced; red X indicates when CSF was sampled but had insufficient cfDNA for analysis. **(B)(i)** Total cell-free DNA (cfDNA) abundance, **(ii)** tumor fraction, and **(iii)** copy number burden (CNB) were calculated from each CSF sample. **(iv)** The variant allele frequency (VAF) for the IDH1 mutation and the levels of D-2-hydroxyglutarate (D-2-HG, in µM) were quantified at each timepoint. **(C)(i-ii)** Copy number plots were generated for each sample; POD0 and 214 shown, as well as (iii) a fishplot depicting the VAFs of mutations from POD0 to 214. **(D)** Observations and questions were raised by patient 138’s data.

CSF was obtained from the patient’s EVD prior to and after resection. cfDNA abundance increased from 31.81 ng/mL to 50.22 ng/mL, while tumor fraction decreased 0.198 to 0.042 (**Fig. 5Bi-ii**). Tumor fraction then increased by 2.87x to 0.121 by suspected progression on POD214 (**Fig. 5Bii)**. CNB decreased and remained below the limit of quantification after resection through POD214 (**Fig. 5Biii**). IDH1 VAF decreased with resection prior to increasing at the time of suspected radiographic progression (POD214), consistent with D-2-HG increasing from 0.4 to 1.02 µM (**Fig. 5Biv**). Chromosome 6p gain and 6q loss, 11q gain, and 13q loss, all common in astrocytoma, IDH-mutant, were identified at POD0, although almost no copy number alterations could be detected by POD214 (**Fig. 5Ci-ii**). TP53 VAF decreased with resection and increased by POD214, along with the emergence of a GNAS mutation (**Fig. 5Ciii)**.

Patient 138 yielded findings reminiscent of those seen in prior patients: 1) as with patient 24, tumor fraction may increase with suspected disease progression (**Fig. 5Bii)**; 2) as with patient 98 (**Fig. 4Biv**), IDH VAF may correlate with changes in D-2-HG (**Fig. 5Biv)**, and 3) as with patient 79 (**Fig. 3Bi)**, resection may transiently increase cfDNA abundance in early post-operative samples (**Fig. 5Bi)**. A remaining question was the threshold at which to call disease progression based on cfDNA.

## DISCUSSION

In this five-patient case series, we report our initial experience with longitudinal intracranial CSF for cfDNA analysis in patients undergoing treatment for glioblastoma or astrocytoma, IDH mutant, grade 4. Although preliminary, our findings provoke the following hypotheses to be further tested: (1) Tumor fraction may increase with suspected disease progression, (2) cfDNA abundance is variable across samples, but may transiently increase post-operatively and with pseudoprogression, (3) changes in mutational variants and their allelic frequencies may be seen within individual patients via longitudinal CSF sampling, (4) CSF D-2-HG levels may correlate with changes in IDH1 cfDNA, despite incongruent radiographic findings, and (5) CNB decreases below the limit of quantification during treatment with current techniques. Key questions included whether pseudoprogression could increase cfDNA abundance and importantly, what is the required threshold of change in each cfDNA measure to call disease progression.

CfDNA burden in other cancer types has been shown to increase with recent resection or treatment, likely due to disruption of the tissue resulting in release of cfDNA^40–42^. Indeed, after resection, CSF cfDNA burden transiently increased for each patient where CSF was obtained prior to and after resection, and remained elevated even by POD21 in patient 78. As resection increases surgical debris that also has its own cell-free DNA, it is possible that ongoing surgical debris may overestimate the relative decrease in tumor fraction based on the ratio of tumor-versus-non-tumor cfDNA. This points to the importance of obtaining a sample immediately prior to chemoradiation, when surgical debris has decreased. Moreover, while some post-resection cfDNA may be attributable to debris, it remains to be established in gliomas whether post-resection cfDNA levels correlate with patient prognosis, as has been identified in other cancer types^43, 44^.

In patient 79, new variants were detected on POD2 after resection. Prior studies, including in glioma, reported that numerous mutations were detected in CSF or plasma, but not in the original tumor tissue^27, 45^. When CSF is obtained prior to surgical resection, it may only contact one portion of the tumor, if at all, depending on the anatomical location. However, resection can then increase CSF contact with new areas of the infiltrative tumor margin as compared to CSF obtained prior to resection^28, 46^. As such, it may be helpful to obtain both a pre-resection and post-resection/pre-chemoradiation sample to determine the full spectrum of detectable tumor mutations for longitudinal monitoring. The latter post-resection sample may then serve as the baseline sample for monitoring disease burden and treatment response.

Prior studies have demonstrated that tumor contact with CSF, as well as tumor size and grade, are key factors for detection of tumor cfDNA in CSF^28, 31, 32, 46^. One study in 21 medulloblastomas, ependymomas, and high-grade gliomas demonstrated that CSF cfDNA could be detected in tumors with CSF contact, whereas no cfDNA was found in tumors that were distant from CSF^28^. Multiple prior studies in other cancer types have noted the superiority of proximal fluids for cfDNA detection^23, 25^. As such, use of CSF access devices in a resection cavity, as performed in our study, is a particularly attractive option for longitudinal CSF acquisition, allowing for close access to the source of the cfDNA. As most gliomas recur around the previous resection cavity^47, 48^, disease progression should be detectable. However, this needs to be demonstrated in a larger cohort of patients.

Our observational case series is intended for hypothesis generation and has numerous limitations. Pre-chemoradiation CSF was not always obtained. CSF cfDNA was also not analyzable in 5/35 samples and CSF cfDNA was not always abundant enough to allow for NGS at each timepoint, hence only LP-WGS could be performed, resulting in missing VAF data. To minimize false positives in CNB calls, intracranial non-tumor control CSF samples will need to be evaluated. Identification of thresholds of change indicative of changes in disease burden will be needed to determine whether changes in cfDNA-associated measures could adjudicate radiographic progression versus treatment effect or predict disease recurrence^49–52^. When available, multiple independent -omics in the same CSF sample, (e.g., D-2-HG) may increase confidence in interpreting cfDNA changes.

## CONCLUSION

Longitudinal acquisition of intracranial CSF cfDNA is feasible and can generate hypotheses regarding the impact of treatment and progression on cfDNA throughout a patient’s disease course. Further studies are needed to determine the thresholds of change in cfDNA metrics that may correlate with changes in disease burden.

## Data Availability

All data produced in the present study are available upon request to the author.

## FUNDING

CRC was supported by the National Institute of Health T32GM145408. AMC was supported by NIH T32GM00868. TCB, TJK, SHK, and JEP were supported by NINDS R61 NS122096. TCB was supported by the Mayo Clinic Center for Individualized Medicine and CCaTS award UL1TR002377, the American Brain Tumor Association, Brains Together for the Cure, Humor to fight the Tumor, and Lucius & Terrie McKelvey.

## AUTHORSHIP

- Study design and conception: TCB, CRC, LPC, AMC, SJ, PD
- Sample acquisition: CLN, KMA, NC, IJT, NM, TCB
- Sample processing and extraction: CRC, WM, SI, KG, ABZ, MDH, AEW
- Analyzed data: CRC, TCB, XD, CD, TJK, RBJ, PD, SJ
- Critically reviewed manuscript: all authors

## DATA AVAILABILITY

All data are available as supplementary files.

## ETHICS

This study was approved by the Mayo Clinic Institutional Review Board and all participants provided their consent to participate in this study. This study was performed in accordance with the Declaration of Helsinki.

## CONSENT

All participants have provided consent for publication.

## ACKNOWLEDGMENTS

We thank our patients and their families for their participation in this study. We thank the Mayo Clinic Neurosurgery Clinical Research, neurosurgery residents, and surgical team for their technical and research support.

## CONFLICT OF INTEREST

Xiaoxi Dong, Chao Dai, Wei Mo, Pan Du, and Shidong Jia report employment at Predicine, Inc. Wei Mo, Pan Du, and Shidong Jia also report stock and other ownership interests in Predicine, Inc.

## Notes

### Clinical Trial

NCT04692337

