## Supplemental figure and methods for "Longitudinal glioma monitoring via cerebrospinal fluid cell-free DNA: one patient at a time"

**Supplementary Figure**

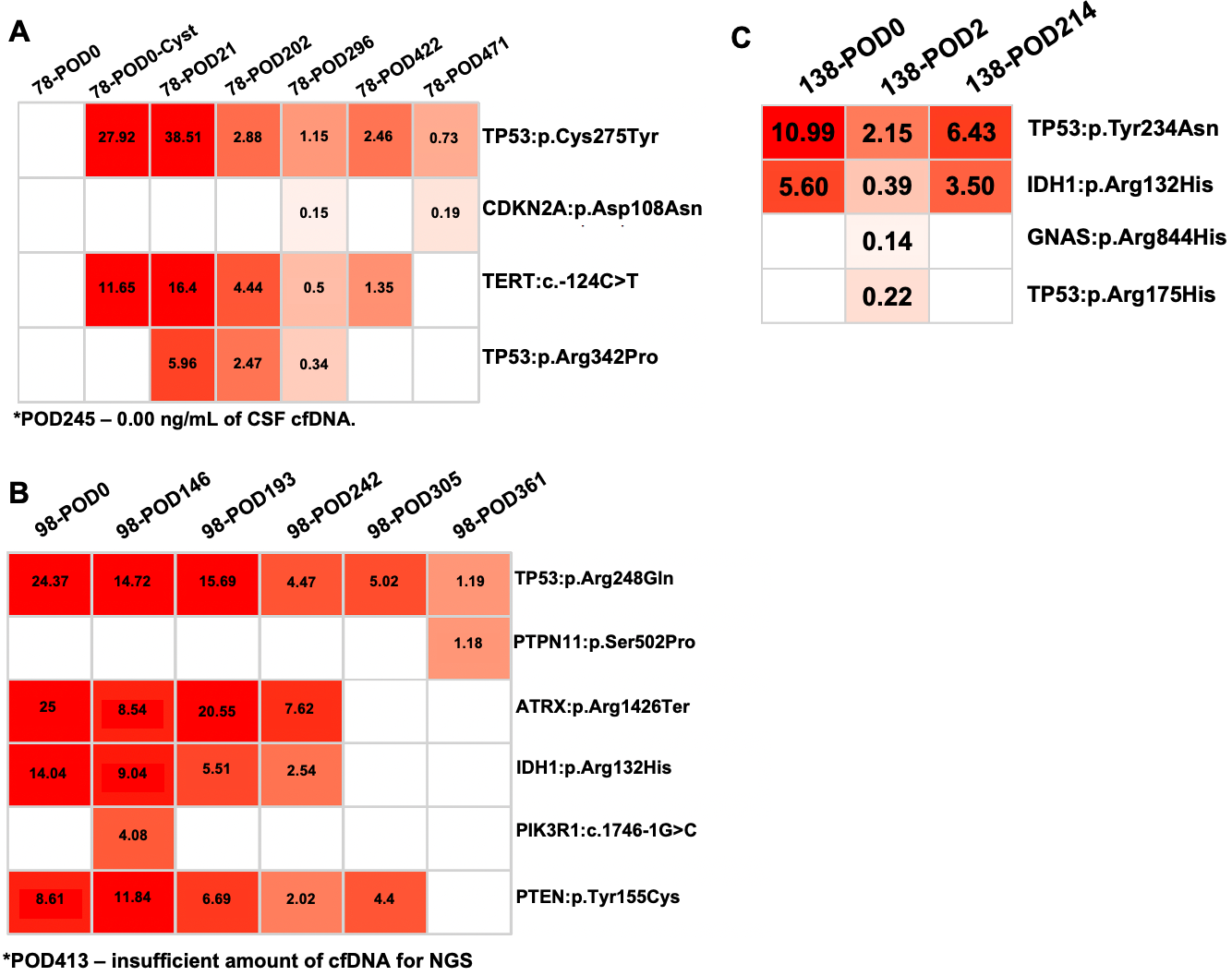

**Supplementary Figure 1. Variant allele frequencies from longitudinal Next-Generation Sequencing in patients 78, 98, and 138.** The variant allele frequencies (VAFs) are shown for longitudinal samples from patients (A) 78, (B) 98, and (C) 138 that underwent Next-Generation Sequencing.

**Supplementary Methods**

**cfDNA extraction**

Cell-free DNA (cfDNA) was extracted from CSF samples using the QIAamp circulating nucleic acid kit (Qiagen, Hilden, Germany). Quantity and quality of the purified cfDNA were checked using a Qubit fluorimeter (ThermoFisher Scientific, Waltham, Massachusetts, USA) and Bioanalyzer 2100 (Agilent Technologies, California, USA).

**Library preparation, hybrid capture and sequencing**

Up to 30 ng of extracted cfDNA were then processed for library construction including end-repair dA-tailing and adapter ligation. Ligated library fragments with appropriate adapters were amplified via PCR. The amplified DNA libraries were then further checked using a Bioanalyzer 2100 and samples with sufficient yield were advanced to hybrid capture. Hybrid capture was conducted using Biotin labelled DNA probes. In brief, each library was hybridized overnight with a Predicine NGS panel and paramagnetic beads. The unbound fragments were washed away, and the enriched fragments were amplified via PCR amplification. The purified product was checked on a Bioanalyzer 2100 and then loaded into an Illumina NovaSeq 6000 (San Diego, CA, USA) for NGS sequencing with paired-end 2x150bp sequencing kits.

**Analyses of NGS data from cfDNA**

Raw sequencing data was processed by aligning to hg19, followed by somatic variant calling and filtering. Details of the methods including ctDNA fraction calculation was described in a previous publication by Davis et al., *Clinical Cancer Research,* 2023.

**Tumor Fraction inferred from LP-WGS CNV**

We developed CNV-based tumor fraction estimation implemented by Expectation-Maximization (EM) algorithm. EM estimates tumor fraction from copy number loss genes, with an assumption that heterozygous (HETD) genes and homozygous (HOMD) genes harbor the same cancer clone. Our EM-based tumor fraction estimation and hidden Markov model (HMM) based tumor fraction estimation (ichorCNA, from Adalsteinsson et al., *Nature Communications*, 2017) are highly consistent.

**Copy number burden analysis using LP-WGS sequencing.**

PredicineSCORE low-pass whole genome sequencing (LP-WGS) with an overall average coverage of 3x was performed on patient samples. Firstly, each 1Mb segment CNV was normalized by GC content and mappability, followed with normalization based on the average of segment CNV profile of normal samples [ichorCNA algorithm, Adalsteinsson et al., *Nature Communications*, 2017]. We further quantified arm-level CNV deviation as the average of segment CNVs across each chromosome arm, and calculated CNB score as the sum of absolute z-score of arm-level CNV deviation, followed with log2 transformation, where higher/lower CNB score indicates higher/lower CNV abnormality compared with normal background. The CNB score cutoff was defined as three standard deviations away from the population mean of normal sample CNB scores.
